# Induction of robust neutralizing antibodies against the COVID-19 Delta variant with ChAdOx1 nCoV-19 or BNT162b2 as a booster following a primary vaccination series with CoronaVac

**DOI:** 10.1101/2021.09.25.21264099

**Authors:** Samadhi Patamatamkul, Sutthiwan Thammawat, Benjaporn Buranrat

## Abstract

We determined the antibody response among CoronaVac vaccinees who were boosted with either BNT162b2 or ChAdOx1 nCoV-19. Increase in anti-S-RBD antibodies and surrogate neutralizing antibodies against the Delta variant was higher in BNT162b2 recipients than in ChAdOx1 nCoV-19 recipients. High proportions of reactogenicity were seen for both booster regimens.

---

As the COVID-19 Delta variant spreads globally, COVID-19 breakthrough infections are increasing despite primary COVID-19 vaccination being completed. In Thailand, most healthcare personnel have been vaccinated with an inactivated virus vaccine, CoronaVac (Sinovac Biotech, Beijing, China). A recent viral neutralization test (VNT) of CoronaVac vaccinees in the Thai population demonstrated markedly reduced titers against the Delta variant as compared to convalescent sera [1]. Therefore, a booster dose of either ChAdOx1 nCoV-19 or BNT162b2 was voluntarily administered to healthcare personnel in Thailand in July–August 2021 as per national policy. Here, we examined the immunogenicity and reactogenicity among healthcare personnel before and after CoronaVac and the third booster dose.

We prospectively enrolled 41 healthcare personnel from Suddhavej Hospital, Mahasarakham University, Mahasarakham, Thailand, for longitudinal collection of blood samples (pre-vaccination, post-second CoronaVac, pre-booster, post-booster) during April to August 2021. All participants were seronegative for anti-S-RBD (spike protein receptor-binding domain) and anti-nucleocapsid antibodies before COVID-19 vaccination, and had not been previously diagnosed with COVID-19 infection.

We used Elecsys Anti-SARS-CoV-2□S (Roche Diagnostics GmbH, Mannheim, Germany) and a cPass™ SARS-CoV-2 NAbs Detection Kit as a surrogate VNT (sVNT) (GenScript, Piscataway, NJ, USA) to examine immunogenicity. The sVNT uses a SARS-CoV-2 spike protein (RBD, L452R, T478K, Avi & His Tag)-HRP reagent, which is specific for the Delta variant S-RBD. The sVNT protocol followed the manufacturer’s instructions, with a 30% inhibition cutoff described elsewhere [2].

Of the 41 participants, 23 received BNT162b2 as the booster dose. Age and gender did not differ significantly among the ChAdOx1 nCoV-19 and BNT162b2 recipients: 38 ± 10.26 vs. 32.04 ± 3.42 years (p = 0.188); 66.7% vs. 60.9% female (p = 0.702). One participant per group had pre-existing diabetes mellitus.

The post-second CoronaVac and pre-booster sera anti-S-RBD antibody levels did not differ significantly: 64.72 U/mL (interquartile range [IQR], 22.23–188.86) vs. 106.8 U/mL (IQR, 49.89–151.7) (p = 0.270) for ChAdOx1 nCoV-19, and 37.78 U/mL (IQR, 16.79–73.8) vs. 37.46 U/mL (IQR, 23.39–51.60) (p = 0.932) for BNT162b2. Anti-S-RBD-antibodies were significantly higher after booster vaccination with BNT162b2 rather than with ChAdOx1 nCoV-19: 22,558 U/mL (IQR, 15,956–25,000) vs. 5,159 U/mL (IQR, 3,647.75–9,196.75) (p < 0.001) (Figure 1A).

**Figure 1.**
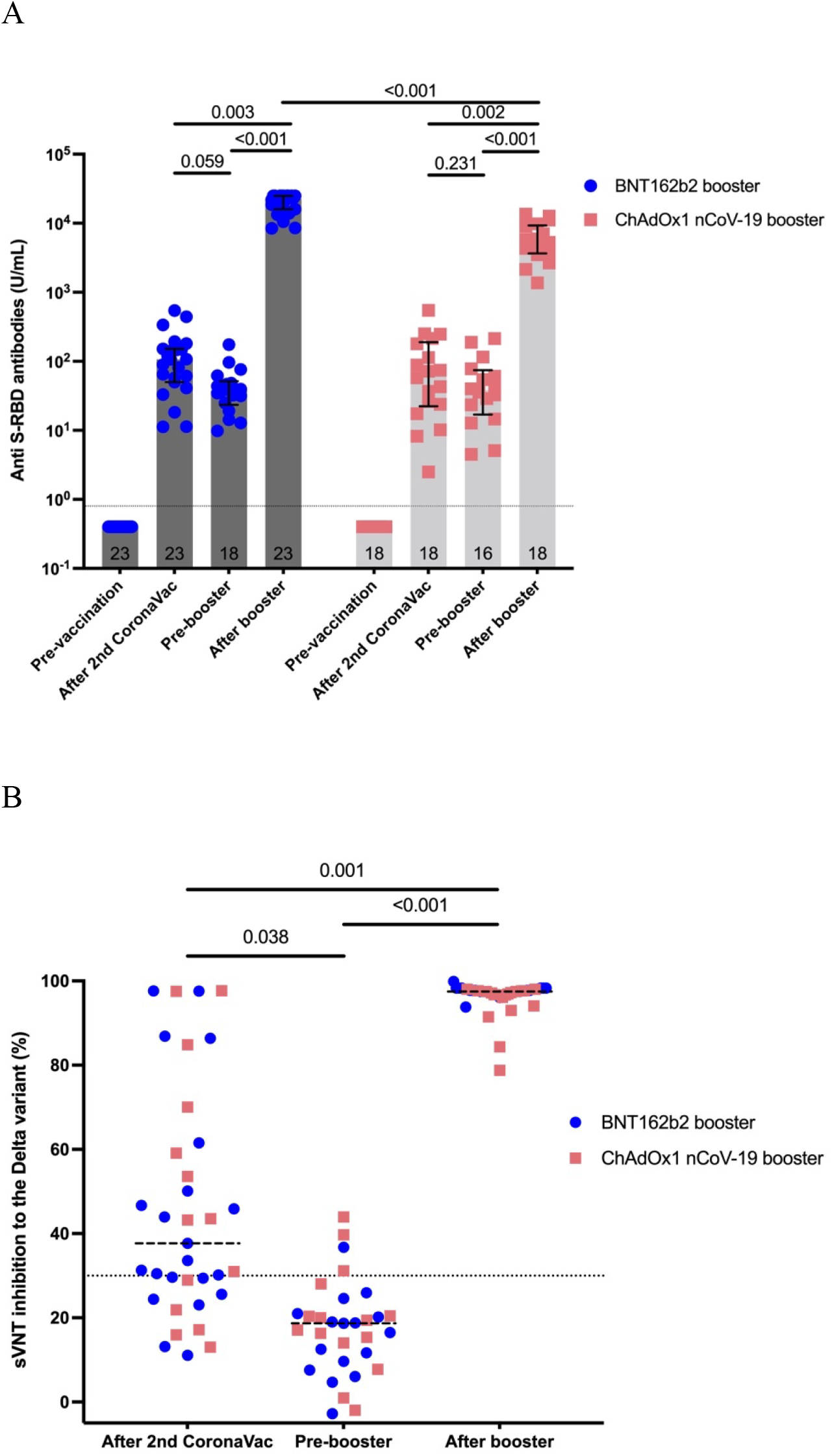
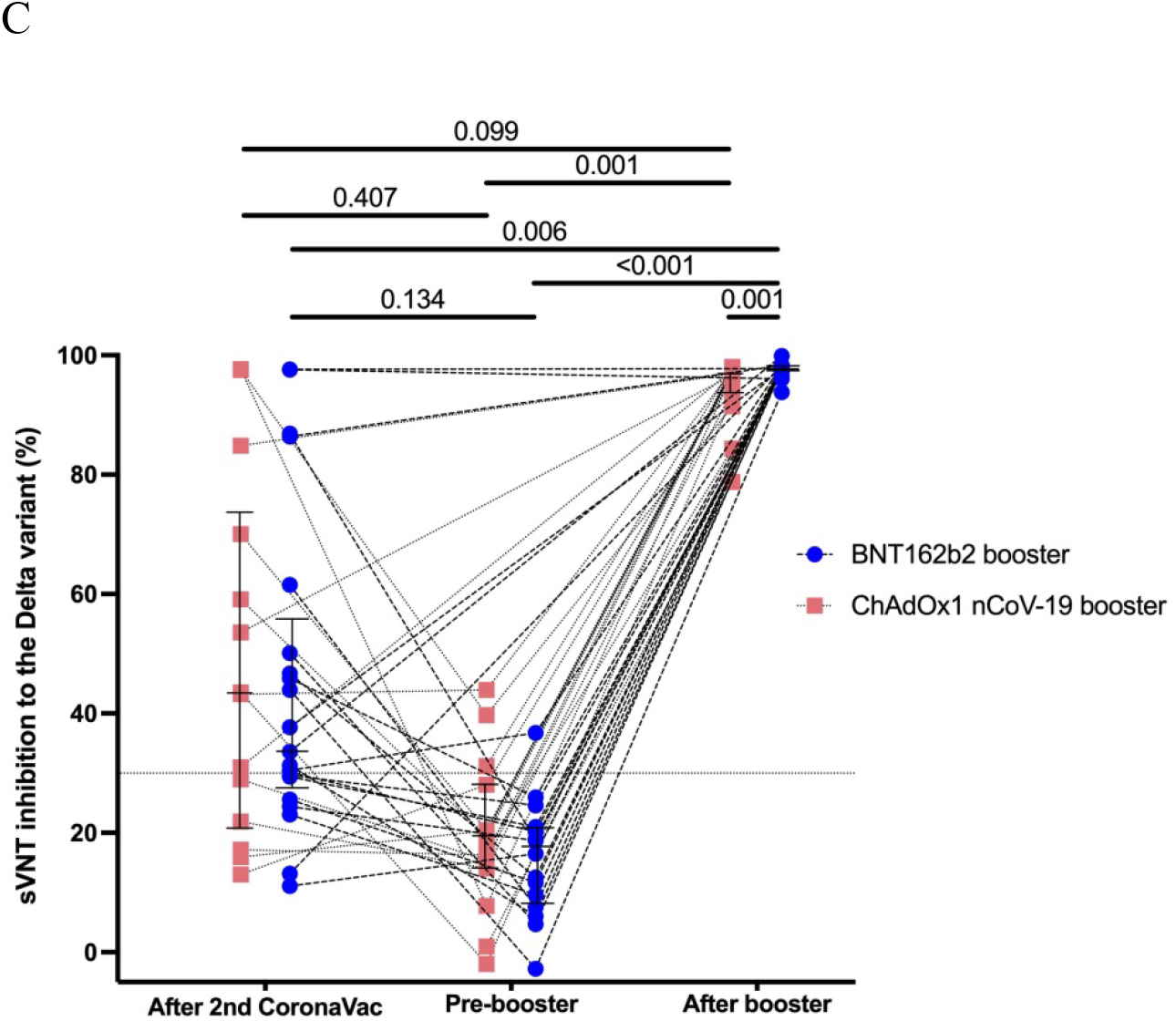
(A) Longitudinal changes in anti-S-RBD antibodies before and after CoronaVac and after booster between BNT162b2 and ChAdOx1 nCoV-19 recipients are shown as separated scatter with bars. The positive cutoff value at ≥0.8 U/mL denotes reactive anti-S-RBD antibodies. Error bars represent the median and IQR. (B) Scatter plots demonstrate longitudinal changes in sVNT in the whole cohort. Dashed lines represent the median. (C) Scatter plots with each replicate connected show longitudinal changes in sVNT between BNT162b2 and ChAdOx1 nCoV-19 recipients; ≥30% inhibition denotes significant inhibition of the Delta variant. P ≤ 0.05 (two-sided) denotes a significant difference using non-parametric Friedman’s two-way analysis with Bonferroni correction for multiple tests. All statistical analyses were performed with SPSS 28 (IBM Corp., Armonk, NY, USA).

Up to 65.7% of CoronaVac vaccinees in the whole cohort achieved sVNT ≥ 30% inhibition, as did 64.3% and 66.7% of ChAdOx1 nCoV-19 and BNT162b2 recipients, respectively. The proportions of participants who maintained pre-booster sVNT ≥ 30% inhibition decreased significantly after the second CoronaVac shot: 65.7% (95% confidence interval [CI], 49.1–79.2) to 12.9% (95% CI, 4.5–29.5) (p < 0.001). After the booster shot, this increased to 100% (95% CI, 89.8–100) (p < 0.001). The median sVNT from after the second CoronaVac to before the booster shot significantly declined: 37.67% (IQR, 25.58–61.54) to 18.71% (IQR, 9.66–20.98). It increased after boosting to 97.52% (IQR, 96.72–98.03) (Figure 1B). BNT162b2 recipients had higher sVNT than ChAdOx1 nCoV-19 recipients: 97.76% (IQR, 97.5–98.29) vs. 97.02% (IQR, 93.8–97.64), respectively (p < 0.001; Figure 1C).

Anti-S-RBD antibodies correlated with the sVNT value (Spearman correlation coefficient of 0.82; 95% CI, 0.75–0.88) (p < 0.001). At anti-S-RBD antibody levels of ≥ 583 U/mL, the sensitivity, specificity, positive predictive value, and negative predictive value for sVNT ≥ 68% were 83.1%, 100%, 100%, and 85.5%, respectively.

The median time between collection of post-second CoronaVac sera and pre-booster sera was not significantly different between the two groups: 4 weeks (IQR, 3–5.25) vs. 4 weeks (IQR, 3–4) for ChAdOx1 nCoV-19, and 11 weeks (IQR, 10–12) vs. 12 weeks (IQR, 11.75–12) for BNT162b2. The median time between the booster dose and serum collection was 3 weeks (IQR, 2.75–3) vs. 2 weeks (IQR, 2–2), respectively (p < 0.001).

All participants had at least one symptom after the booster (Table 1). Up to 92.5% of participants used acetaminophen for relieving symptoms.

**Table 1.**
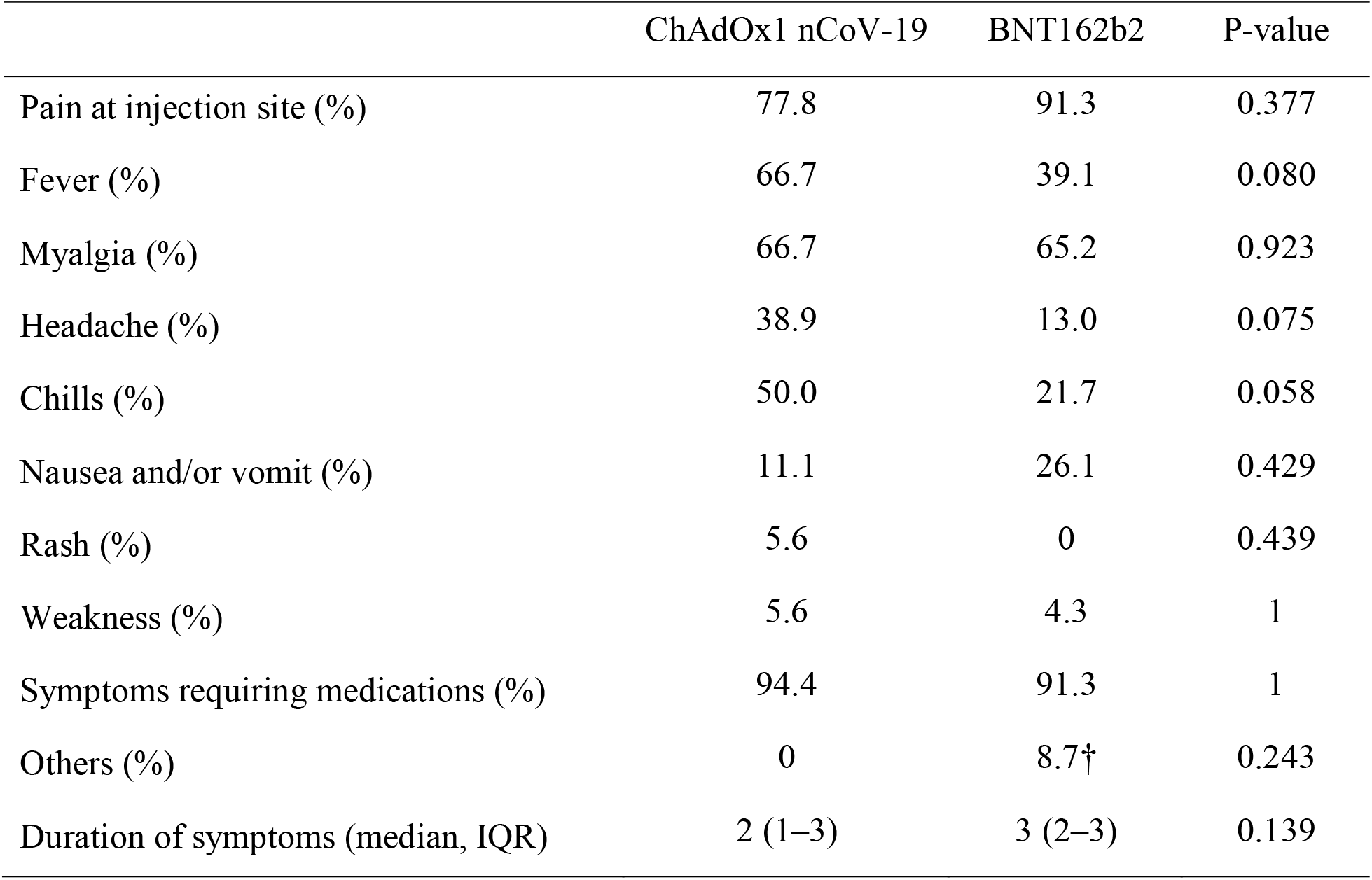
† One participant had peripheral bilateral numbness of the hands, achieving recovery after 1-week gabapentin; another had acute tonsillitis requiring antibiotics. Proportions of reactogenicity in each group

This is the first prospective study of a booster dose with viral vector or mRNA vaccine after CoronaVac vaccination in the Thai population. Our findings demonstrate a robust antibody response in terms of both anti-S-RBD antibodies and sVNT after boosting with ChAdOx1 nCoV-19, and even more so with BNT162b2. All participants achieved more than 68% inhibition against the Delta variant, which is the cutoff for high-titer neutralizing antibodies according to the US-FDA. Our findings are concordant with the mouse model, where a heterologous vaccine booster with either viral vector or mRNA vaccine after completion of the primary regimen with an inactivated vaccine significantly increased the levels of neutralizing antibodies [3].

With regard to CoronaVac durability, we found that only 12.9% maintained sVNT at above the cutoff after 3 months. Our findings are concordant with the previous study regarding the persistence of anti-spike antibodies with decreasing neutralizing effect [4].

We found overall higher reactogenicity in our cohort than previously reported with BNT162b2 or ChAdOx1 nCoV-19 [5].

In conclusion, after CoronaVac primary vaccination, both BNT162b2- and ChAdOx1 nCoV-19-boosted recipients demonstrated consistently high neutralizing antibodies against the Delta variant. BNT162b2 may be a more favorable booster, with significantly higher neutralizing antibodies, which may correlate with better clinical effectiveness.

## Data Availability

Datasets are available upon request.

## Funding source

This work was supported by the Faculty of Medicine, Mahasarakham University.

## Conflicts of interests

The authors declare no conflicts of interest.

## Acknowledgements

We thank all the laboratory technicians who helped with the sample collection throughout the study.

## Ethical approval

The Mahasarakham University Ethics Committee approved the study. Informed consent was obtained before blood collection.

## Authors’ contributions

SP: Designed the study, interpreted the findings, and wrote the paper.

BB: Performed the laboratory experiments and interpreted the findings.

ST: Performed the laboratory experiments and interpreted the findings.

## Notes

### Competing Interest Statement

The authors have declared no competing interest.

## References

1. Vacharathit V, Aiewsakun P, Manopwisedjaroen S, Srisaowakarn C, Laopanupong T, Ludowyke N, et al. CoronaVac induces lower neutralising activity against variants of concern than natural infection. Lancet Infect Dis 2021. https://doi.org/10.1016/S1473-3099(21)00568-5.

2. Tan CW, Chia WN, Qin X, Liu P, Chen MI-C, Tiu C, et al. A SARS-CoV-2 surrogate virus neutralization test based on antibody-mediated blockage of ACE2-spike protein-protein interaction. Nat Biotechnol 2020;38:1073–8. https://doi.org/10.1038/s41587-020-0631-z.

3. Zhang J, He Q, An C, Mao Q, Gao F, Bian L, et al. Boosting with heterologous vaccines effectively improves protective immune responses of the inactivated SARS-CoV-2 vaccine. Emerg Microbes Infect 2021;10:1598–608. https://doi.org/10.1080/22221751.2021.1957401.

4. Widge AT, Rouphael NG, Jackson LA, Anderson EJ, Roberts PC, Makhene M, et al. Durability of Responses after SARS-CoV-2 mRNA-1273 Vaccination. N Engl J Med 2021;384:80–2. https://doi.org/10.1056/NEJMc2032195.

5. Hillus D, Schwarz T, Tober-Lau P, Vanshylla K, Hastor H, Thibeault C, et al. Safety, reactogenicity, and immunogenicity of homologous and heterologous prime-boost immunisation with ChAdOx1 nCoV-19 and BNT162b2: a prospective cohort study. Lancet Respir Med 2021. https://doi.org/10.1016/S2213-2600(21)00357-X.

